# Coronary Assessment in Heart Failure within a Safety-Net Setting: Disparities and Outcomes

**DOI:** 10.1101/2023.07.06.23292331

**Authors:** Matthew S. Durstenfeld, Anjali Thakkar, Yifei Ma, Lucas S. Zier, Jonathan D. Davis, Priscilla Y. Hsue

## Abstract

**Background:** Though ischemic cardiomyopathy is the leading cause of heart failure (HF), most patients do not undergo coronary assessment after heart failure diagnosis. In a safety-net population, referral patterns have not been studied, and it is unknown whether coronary assessment is associated with improved HF outcomes.

**Methods:** Using an electronic health record cohort of all individuals with HF within San Francisco Health Network from 2001-2019, we identified factors associated with completion of coronary assessment (invasive coronary angiography, nuclear stress, or coronary computed tomographic angiography). Then we emulated a randomized clinical trial of elective coronary assessment with outcomes of all-cause mortality and a composite outcome of mortality and emergent angiography. We used propensity scores to account for differences between groups. We used national death records to improve ascertainment of mortality.

**Results:** Among 14,829 individuals with HF (median 62 years old, 5,855 [40%] women), 3,987 (26.9%) ever completed coronary assessment, with 2,467 (18.5%) assessed out of 13,301 with unknown CAD status at HF diagnosis. Women and older individuals were less likely to complete coronary assessment, with differences by race/ethnicity, medical history, substance use, housing, and echocardiographic findings. Among 5,972 eligible for inclusion in the “target trial,” 627 underwent early elective coronary assessment and 5,345 did not. Coronary assessment was associated with lower mortality (HR 0.84; 95% CI 0.72-0.97; p=0.025), reduced risk of the composite outcome, higher rates of revascularization, and higher use of medical therapy.

**Conclusions:** In a safety-net population, disparities in coronary assessment after HF diagnosis are not fully explained by CAD risk factors. Our target trial emulation suggests coronary assessment is associated with improved HF outcomes possibly related to higher rates of revascularization and GDMT use, but with low certainty that this is finding is not attributable to unmeasured confounding.

Graphical Abstract:

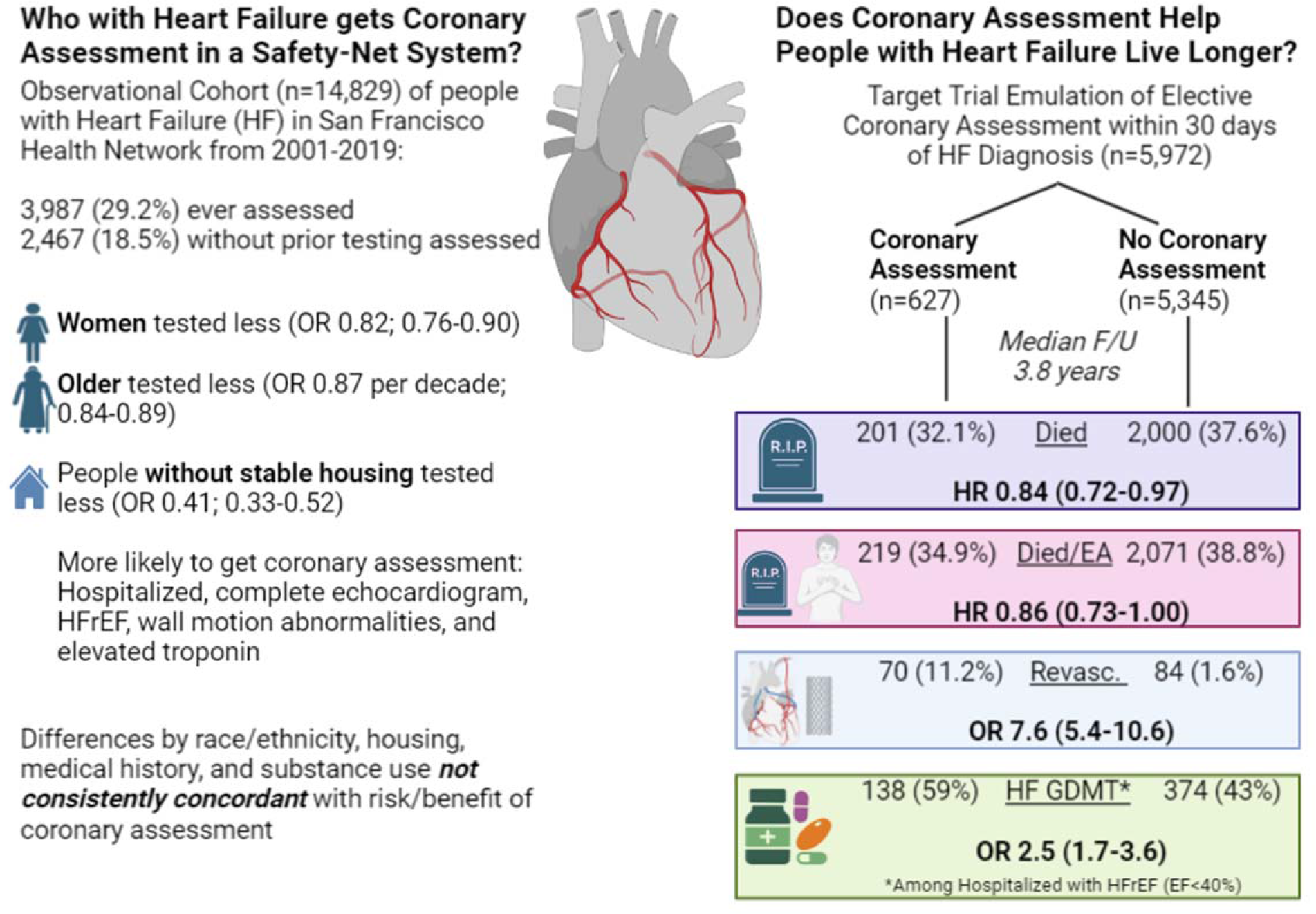

## What is Known

A low proportion of individuals undergo coronary assessment after heart failure diagnosis.

Coronary assessment is associated with improved outcomes in observational studies but has not been studied in a randomized controlled trial.

## What the Study Adds

Within a safety-net population, there are significant disparities in who completes coronary assessment after heart failure diagnosis that are not explained by risk factors for ischemic cardiomyopathy.

In a target trial to emulate a randomized clinical trial, elective coronary assessment was associated with improved survival, revascularization, and higher rates of prior GDMT use.

Although we used rigorous causal inference methods, answers to this important clinical question remain inconclusive without a randomized clinical trial.

## Introduction

Heart failure (HF) is a major cause of morbidity and mortality with >1,000,000 new cases per year in the United States^1^ and disproportionately affects those who identify as Black/African American.^2^ Ischemic cardiomyopathy attributable to obstructive coronary artery disease (CAD) accounts for 60-70% of HF cases.^3, 4^ According to the 2022 AHA/ACC/HFSA Guideline for the Management of Heart Failure, “*In patients with HF, an evaluation for possible ischemic heart disease can be useful to identify the cause and guide management”* (Level 2a recommendation).^5^ Importantly, the recommendation applies to both HF with a reduced ejection fraction (HFrEF) and HF with a preserved ejection fraction (HFpEF), which is also commonly caused by coronary artery disease. This is partly based on the 10-year follow up of the STICH trial (STICHES), which found that coronary artery bypass graft (CABG) surgery added to medical therapy had a mortality benefit for patients with HFrEF due to ischemic cardiomyopathy. REVIVED-BCIS2, which randomized patients with ischemic HFrEF and evidence of viability to revascularization with percutaneous coronary intervention, did not find an intermediate term benefit.^7^ This has led some cardiologists to question routine coronary assessment for patients with HF.

To our knowledge, there are no randomized clinical trials of coronary assessment strategies among patients with HF. Observational studies suggest that coronary assessment during HF hospitalization or within 2 weeks of diagnosis is associated with higher use of medical therapy and revascularization, and with reduced mortality and rehospitalization.^8, 9^ Despite the evidence, most commercially-insured patients with HF do not undergo coronary assessment within 90 days, with disparities by county, patient demographics, and co-management by a cardiologist.^10^ Women and persons of Black race are less likely to be referred for coronary assessment in other settings.^11^ None of the published studies have examined coronary assessment in a safety-net setting, and most do not adequately account for selection effects for coronary assessment, exclude patients with acute coronary syndromes, or align timing and eligibility in an effort to emulate a randomized clinical trial to answer these questions.

We therefore designed this study to examine whether there are differences in who undergoes coronary assessment among those with HF within a safety-net setting, and secondly whether elective coronary assessment early after HF diagnosis among those without known CAD is associated with clinical outcomes including mortality and a combined outcome of mortality and emergent coronary angiography. We hypothesized that there would be disparities in coronary assessment by patient demographics not explained by CAD risk factors. Our second hypothesis was that early coronary assessment would be associated with lower mortality and lower risk of subsequent emergent angiography among those without prior coronary assessment or indication for emergent coronary angiography.

## Methods

### Study Design and Participants

We developed an electronic health record (EHR) cohort of all individuals with HF by ICD-9 or ICD-10 code who received care within San Francisco’s municipal health system, the San Francisco Health Network (SFHN), during 2001-2019. HF was defined as ICD-9 codes: 428, 428.0, 428.1, 428.2X, 428.3X, 428.4, 428.9, 402.01, 402.11, 402.91, 404.01, 404.03, 404.11, 404.13, 404.91, 404.93 and ICD-10 codes: I50.1, I50.20, I50.21, I50.22, I50.23, I50.30, I50.31, I50.32, I50.33, I50.40, I50.41, I50.42, I50.43, I50.9, I11.0) coded during an outpatient or inpatient healthcare encounter. Patients were included from January 1, 2001-August 1, 2019, and last follow up was December 31, 2019.

### Exposures

Left heart catheterization with invasive coronary angiography, exercise and pharmacologic nuclear stress tests, and coronary computed tomographic angiography (CCTA) were considered as coronary assessments. For those with coronary assessments and echocardiograms, full texts of reports were extracted, including a look back period to 1999. We categorized the results using structured text extraction with manual review for refining the structured test extraction and verifying quality control. After extraction, all reports were manually reviewed for accuracy by the first author.

### Outcomes

The two primary outcomes were all-cause mortality and a composite outcome of all-cause mortality and emergent coronary angiography for STEMI, NSTEMI, cardiogenic shock, ventricular arrhythmias, and cardiac arrest ascertained by manual review of all cardiac catheterization reports by the first author. Emergent angiography performed at other sites was not included. Patients were linked with Social Security Death Index/National Death Index (SSDI/NDI) records for all cause mortality by name, birth date, and social security number (if available). For patients who could not be linked, vital status and date of death if deceased were abstracted from the medical records.

### Additional Variables

We extracted past medical history at the time of HF diagnosis using ICD codes. Psychiatric comorbidities were not measured. Using the first available echocardiogram concurrent with HF, we classified as HFrEF those with left ventricular ejection fraction (LVEF) <40%; if LVEF was not reported (as was common in earlier years of the study), we classified those with at least moderately reduced left ventricular (LV) function on qualitative assessment as HFrEF; we classified those with LVEF≥41% if reported or mild-to-moderate LV systolic dysfunction or less on qualitative assessment if quantitative LV was not reported as HFpEF. Medical records were manually reviewed for all individuals with a cardiac catheterization report with obstructive CAD (one or more vessels >80%) to ascertain revascularization outcomes. Among those hospitalized for HF, we extracted ambulatory prescription records for before and after the index hospitalization.

### Statistical Analysis: Factors Associated with Coronary Assessment

We estimated the association between ever undergoing coronary assessment and baseline variables with adjustment for age, sex, race/ethnicity, documented unstable housing, diabetes, hypertension, chronic kidney disease, HIV, tobacco, and other substance use, having completed an echocardiogram, HFrEF, and presence of regional wall motion abnormalities using logistic regression.

### Target Trial Emulation of a Hypothetical Randomized Trial of Coronary Assessment

To estimate the effect of coronary assessment on the two outcomes, we emulated a hypothetical randomized clinical trial (“target trial”) of patients with incident HF without prior coronary assessment or known CAD in which we would randomize patients to undergo coronary assessment within 30 days of HF diagnosis.

### Target Trial Inclusion and Exclusion Criteria

We included patients age <80 years old with incident HF starting in the second year of the study period (2002) so we could exclude those with prevalent heart failure during the first year of the study (2001). We also excluded those with prior coronary assessment/known CAD (with a 3-year look back period to 1999), metastatic cancer, advanced cirrhosis, and initial presentation with an acute coronary syndrome (STEMI or NSTEMI), ventricular arrhythmias, cardiac arrest, and those with concurrent endocarditis, severe aortic stenosis or regurgitation, and severe mitral stenosis where there would not be equipoise to randomize patients to no coronary assessment, and those who did not complete an echocardiogram.

### Target Trial Alignment of Eligibility and Follow Up Time

We allowed a 30-day grace period for coronary assessment after diagnosis with heart failure to minimize immortal time bias. We set the start of follow up and eligibility to the date of heart failure diagnosis. Patients were censored at death using the SSDI/NDI date of death, at the end of the SSDI/NDI search for those matched and alive (December 31, 2018 (n=267) or December 31, 2019 (n=1396)), and EHR last contact date and vital status for those who could not be matched (n=1,647).

### Statistical Analysis

We generated a logistic propensity score model for coronary assessment including the restricted cubic spline of age, sex, race/ethnicity, unstable housing, medical history, substance use, hospitalization at the time of diagnosis with heart failure, EF category concurrent or preceding HF, and diagnosis year, all of which are variables known prior to the treatment “assignment.” We assessed the proportion tested by propensity score quintile, balance of covariates across propensity score quintiles, and goodness-of-fit using the Hosmer-Lemshow test. We used Cox proportional hazards models to estimate the hazard ratio for mortality and for the composite outcome by coronary assessment status adjusted for age, sex, the restricted cubic spline of the propensity score for testing coronary assessment, and HF hospitalization at the time of diagnosis. Because the proportional hazards assumption was violated with non-parallel log-log plots of survival and test of the scaled Schoenfeld residuals, we incorporated sex by age interaction terms and conducted sensitivity analyses with truncated follow up times (Supplemental Figure).

We conducted subgroup analyses considering differences by sex, HFrEF vs HFpEF, HF hospitalization at diagnosis, and study period dichotomized into 2002-2012 and 2013-2019 by introducing interaction terms. To consider possible mechanisms for improvement in mortality among those who underwent coronary assessment we classified participants based on the results of the testing and conducted an exploratory analysis considering use of goal-directed medical therapy among hospitalized patients for whom prescription records were available. We also considered the role of revascularization.

To check the robustness of our findings to our analytic choices, we conducted sensitivity analyses examining the role of censoring time on our findings and using inverse probability of treatment weighting (IPTW) with dropping the most extreme 5% of weights (those who were tested despite very low propensity score, which suggests that unmeasured factors may be playing a role in the decision to refer the patient) as an alternative analytic strategy. We used the IPTW results to estimate the average treatment effect on mortality at 4 years and overall. As another alternative approach we used propensity matching with 2:1 (untested: tested) nearest neighbor matching based on the Mahalanobis distance again to estimate the average treatment effect on mortality at 4 years and overall.

Although our primary interest was the estimated hazard ratios and confidence intervals, we considered p<0.05 significant for the two primary outcomes, p<0.001 significant for univariate analyses, and p<0.10 significant for interactions. Analyses were performed using STATA version 17.1. IRB approval was granted by the University of California with a waiver of informed consent. Results are reported in accordance with STROBE guidelines.^12^

## Results

### Description of Cohort & Coronary Assessment

The cohort included 14,829 individuals with HF who received care within SFHN from 2001-2019. The median age at diagnosis was 62 (IQR 53, 75). There were 5,855 women (40%), and a high proportion of Black/African American, Asian, and Hispanic/Latino individuals (Table 1). Among those with HF (both HFrEF and HFpEF), 3,987 (26.9%) completed at least one coronary assessment, with 1163 (29.2%) completing their first coronary assessment more than 30 days prior to heart failure diagnosis, 1169 (29.3%) within 30 days before or after heart failure diagnosis, and 1655 (41.5%) more than 30 days after HF diagnosis (Table 2). For comparison, 11,172 (75.4%) ever completed an echocardiogram; 6,289 (42.4%) had at least one echocardiogram within 30 days of diagnosis, and 5,839 (39.4%) had one more than 30 days after diagnosis. Of those tested, nearly three quarters underwent invasive coronary angiography as the initial test, with exercise or pharmacologic nuclear stress among nearly all the rest. About one third each underwent coronary assessment >30 days prior to HF diagnosis, within 30 days, and more than 30 days after diagnosis. There are differences in test type by timing, with nearly 90% assessed within 30 days completing invasive angiography (p<0.001).

**Table 1.** Participant Characteristics by Coronary Assessment.

Legend. P-values were estimated using chi-squared test for categorical variables and t-tests for continuous variables; $p < 0.01$ was considered statistically significant.
|  | Coronary<br>Assessment<br>(n=3,987) | No Coronary<br>Assessment<br>(n=10,842) | Unadjusted<br>p-value |
| --- | --- | --- | --- |
| Age | 60.3 (52.4, 68.7) | 63.2 (53.1, 77.4) | <0.001 |
| Female<sup>a</sup> | 1390 (35.0%) | 4465 (41.7%) | <0.001 |
| **Race/Ethnicity** |  |  |  |
| White | 843 (21.1%) | 3164 (29.2%) | <0.001 |
| Black/African American | 1096 (27.5%) | 3034 (28.0%) |  |
| Asian | 933 (23.4%) | 2243 (20.7%) |  |
| American Indian/Alaskan Native | 46 (1.2%) | 112 (1.0%) |  |
| Native Hawaiian/ Pacific Islander | 12 (0.3%) | 35 (0.3%) |  |
| Hispanic/Latino | 957 (24.0%) | 1851 (17.1%) |  |
| Other/Decline to state | 100 (2.5%) | 403 (3.7%) |  |
| Documented Unstable Housing | 96 (2.4%) | 565 (5.2%) | <0.001 |
| **Past Medical History** |  |  |  |
| Hypertension | 3557 (89.2%) | 8359 (77.1%) | <0.001 |
| Diabetes | 313 (7.9%) | 934 (8.6%) | 0.14 |
| Chronic Kidney Disease | 582 (14.6%) | 1621 (15.0%) | 0.59 |
| HIV | 221 (5.5%) | 685 (6.3%) | 0.08 |
| **Substance Use** |  |  |  |
| Alcohol | 945 (23.7%) | 2222 (20.5%) | <0.001 |
| Tobacco | 2057 (51.6%) | 4046 (37.3%) | <0.001 |
| Cocaine | 629 (15.8%) | 1462 (13.5%) | <0.001 |
| Methamphetamine | 461 (11.6%) | 1301 (12.0%) | 0.47 |
| Opioid | 398 (10.0%) | 1188 (10.9%) | 0.09 |
| HF hospitalization (ever) | 563 (14%) | 1076 (9.9%) | <0.001 |
| **Echocardiographic Parameters** |  |  |  |
| Ever had an echocardiogram | 3,874 (97.2%) | 7,298 (67.4%) | <0.001 |
| Echocardiogram within 30 days of HF diagnosis | 2132 (53.5%) | 4157 (38.3%) | <0.001 |
| LVEF as measured >30 days before diagnosis, % | 48±13 | 51±12 | <0.001 |
| LVEF measured within 30 days of HF diagnosis, % | 38±15 | 43±16 | <0.001 |
| HFrEF<sup>b, c</sup> | 1,567 (40.4%) | 2,123 (29.1%) | <0.001 |
| Regional Wall Motion Abnormalities Reported<sup>c</sup> | 661 (46.5%) | 433 (24.2%) | <0.001 |
| Severe Pulmonary Hypertension<sup>c</sup> | 66 (3.1%) | 117 (2.8%) | 0.53 |
| Estimated Pulmonary Artery Systolic Pressure<sup>c</sup> | 36±15 | 37±15 | 0.17 |
| Severe Aortic Stenosis<sup>c</sup> | 15 (0.7%) | 13 (0.3%) | 0.03 |
| Severe Aortic Regurgitation<sup>c</sup> | 24 (1.1%) | 16 (0.4%) | <0.001 |
| Severe Mitral Stenosis<sup>c</sup> | 6 (0.3%) | 5 (0.1%) | 0.15 |
| Severe Mitral Regurgitation<sup>c</sup> | 188 (8.8%) | 253 (6.1%) | <0.001 |
| Severe Tricuspid Regurgitation<sup>c</sup> | 144 (6.8%) | 402 (9.7%) | <0.001 |

**Table 2.** Type and Timing of Initial Coronary Assessment (n=3,987)

Legend: Number and percentage who underwent each type of testing by test timing category (column percentage except total row which is row percentage). Abbreviations: CCTA=Coronary Computed Tomographic Angiography; HF=Heart Failure.
| Test Type | Testing Completed<br>>30 days<br>prior to HF<br>diagnosis | Testing Completed<br>Within 30<br>days of HF<br>diagnosis | Testing Completed<br>>30 days after<br>HF diagnosis | Total |
| --- | --- | --- | --- | --- |
| Invasive<br>Coronary<br>Angiography | 682 (58.6%) | 1051 (89.9%) | 1107 (66.9%) | 2840 (71.2%) |
| Nuclear Stress | 477 (41.0%) | 114 (9.8%) | 532 (32.2%) | 1123 (28.2%) |
| CCTA | 4 (0.3%) | 4 (0.3%) | 16 (1.0%) | 24 (0.6%) |
| Total | 1163 (29.2%) | 1169 (29.3%) | 1655 (41.5%) | 3,987 |

Excluding those with prior coronary assessment (n=1163), prior percutaneous coronary intervention or CABG performed elsewhere (n=284), only 2,467/13,301 (18.5%) of individuals with unknown CAD status at the time of HF diagnosis underwent coronary assessment concurrent with or after HF diagnosis. Among those with HFrEF, 1,082/3,204 (33.8%) underwent coronary assessment concurrent with or after HF diagnosis. A lower proportion completed coronary assessment in more recent years of the study (Figure 1, p<0.001).

**Figure 1.**
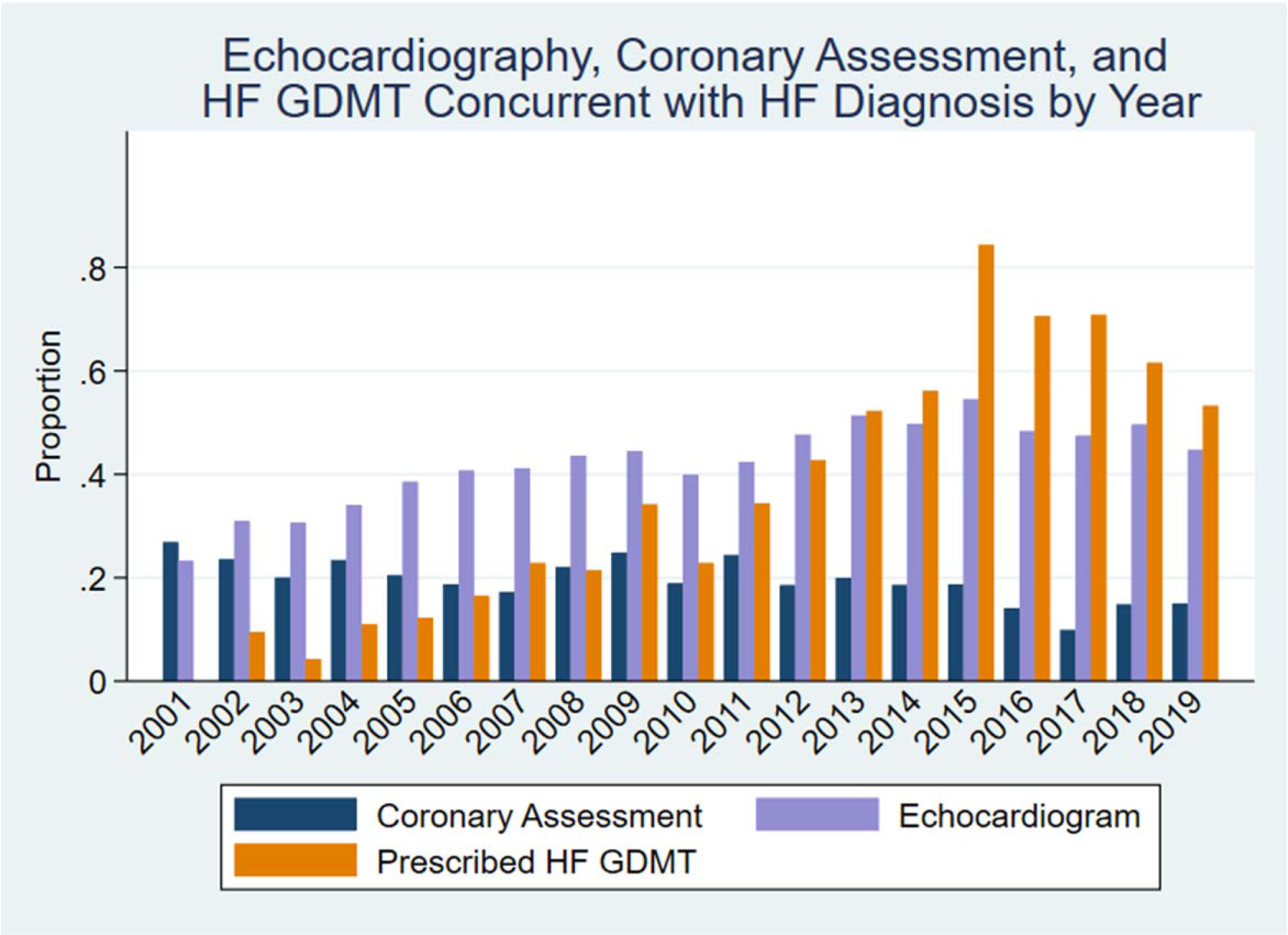
Trends in Echocardiography and Coronary Assessment by Year. The proportion of individuals with incident heart failure who completed coronary assessment (navy) and echocardiogram (lavender) within 30 days of diagnosis and the proportion hospitalized with HFrEF at the time of diagnosis prescribed outpatient GDMT (orange) by year. There was a statistically significant trend for less coronary assessment completed over time and much higher rates of GDMT prescription in the more recent years of the study

### Disparities in Coronary Assessment

In univariate analysis, women and white individuals were less likely to complete coronary assessment (Table 1). A higher proportion who completed coronary assessment had hypertension, alcohol, tobacco, or cocaine use. Individuals who completed testing were more likely to be hospitalized for HF, have completed an echocardiogram concurrent with HF diagnosis, have HFrEF compared to HFpEF, and have regional wall motion abnormalities reported on echocardiogram.

In models adjusted for age, sex, race/ethnicity, housing status, medical history, substance use, hospitalization for heart failure, and ever having an echocardiogram, there were lower odds of completing coronary assessment among those with older age, female sex, and unstable housing (Table 3). Compared to white individuals, Asian and Hispanic/Latino individuals had higher odds of ever completing coronary assessment, but no difference in concurrent assessment. Black/African American individuals, although not less likely to ever have coronary assessment, were much less likely to have their coronaries assessed concurrent with HF diagnosis (OR 0.28; 95% CI 0.11-0.74), with no differences among Hispanic/Latino and Asian individuals compared to white individuals in concurrent coronary assessment. Hypertension and tobacco use were associated with higher odds of coronary assessment, but diabetes, chronic kidney disease, HIV, and methamphetamine use were associated with lower odds of completing coronary assessment. Ever completing an echocardiogram, a crude surrogate measure for completing cardiac testing, was associated with much higher odds of completing coronary assessment; among those who completed an echocardiogram, having HFrEF or regional wall motion abnormalities was associated with higher odds of testing. As expected, presentation with an acute coronary syndrome was associated with much higher odds of coronary assessment.

**Table 3.** Adjusted Odds of Ever Completing Coronary Assessment.

Legend: We found significant differences in who completed coronary assessment by age, sex, race/ethnicity, past medical history, and substance use.
|  | Adjusted Odds Ratio | 95% CI | Adjusted P value | Interpretation |
| --- | --- | --- | --- | --- |
| **Age, per decade** | 0.87 | 0.84, 0.89 | <0.001 | Discordant with ischemic cardiomyopathy risk, but possibly concordant with perceived coronary assessment risk/benefit |
| **Female<sup>a</sup>** | 0.82 | 0.76, 0.90 | <0.001 | As this is adjusted for age, it may reflect sexism in the perception that men have higher risk |
| **Race/Ethnicity** |  |  |  | May reflect structural racism and implicit bias |
| White | Reference |  |  |  |
| Black/African American | 1.06 | 0.94, 1.19 |  |  |
| Asian | 1.53 | 1.36, 1.73 | <0.001 |  |
| American Indian/Alaskan Native | 1.39 | 0.95, 2.02 |  |  |
| Native Hawaiian/ Pacific Islander | 0.75 | 0.38, 1.50 |  |  |
| Hispanic/Latino | 1.79 | 1.58, 2.02 | <0.001 |  |
| Other/Decline to state | 1.70 | 1.29, 2.25 | <0.001 |  |
| **Documented Unstable Housing** | 0.41 | 0.33, 0.52 | <0.001 | May reflect concerns regarding follow up as well as higher burden of substance use and psychiatric illness |
| **Past Medical History** |  |  |  |  |
| Hypertension | 1.85 | 1.64, 2.09 | <0.001 | Concordant |
| Diabetes | 0.75 | 0.65, 0.87 | 0.001 | Discordant |
| Chronic Kidney Disease | 0.86 | 0.77, 0.97 | 0.002 | Discordant with risk of CAD, but concordant with expected risk of testing and benefit of revascularization |
| HIV | 0.78 | 0.67, 0.93 | 0.01 | Discordant |
| Cirrhosis | 0.32 | 0.15, 0.68 | 0.003 | Concordant |
| **Substance Use** |  |  |  |  |
| Alcohol | 0.84 | 0.75, 0.93 | <0.001 | Concordant |
| Tobacco | 1.38 | 1.26, 1.51 | <0.001 | Concordant |
| Cocaine | 1.05 | 0.92, 1.21 |  | N/A |
| Methamphetamine | 0.79 | 0.68, 0.91 | 0.001 | Controversial |
| Opioid | 0.75 | 0.65, 0.86 | <0.001 | Controversial |
| **HF hospitalization** | 1.14 | 1.02, 1.29 | 0.02 | Concordant |
| **Ever had an echocardiogram** | 13.0 | 10.7, 15.9 | <0.001 | Concordant |
| HFrEF (vs HFpEF)<sup>a</sup> | 1.80 | 1.64, 1.97 | <0.001 | Controversial |
| Regional Wall Motion Abnormalities<sup>a</sup> | 2.78 | 2.36, 3.27 | <0.001 | Concordant |
| Troponin I >0.04 ng/mL<sup>b</sup> | 2.24 | 1.78, 2.82 | <0.001 | Concordant |
| STEMI<sup>b</sup> | 100 | 58, 173 | <0.001 | Concordant |
| NSTEMI<sup>b</sup> | 479 | 67, 3436 | <0.001 | Concordant |

### Findings on Coronary Assessment

Among 3,987 individuals who ever underwent coronary assessment, on their first test 1,429 (36.1%) had no CAD or a negative stress test, 855 (21.6%) had nonobstructive CAD, 1,269 (32%) had obstructive CAD or a positive stress test, 322 (8.1%) had evidence of prior revascularization, and 89 (2.3%) had possible ischemia or a nondiagnostic test. Among those who underwent a nuclear stress (n=1,359), 1029 (75.7%) were negative, 91 (6.7%) had possible ischemia, 160 (11.8%) were positive for ischemia, and 79 (5.8%) were nondiagnostic. Among those who underwent invasive coronary angiography (n=3,190), 602 (18.9%) had no CAD, 956 (30.0%) had non-obstructive CAD, 469 (14.7%) had single vessel obstructive CAD, 814 (25.5%) had multivessel obstructive CAD including 329 with obstructive left main disease (≥50%) and 468 with 3 or more vessels with obstructive disease (≥80% as reported visually in angiography report or left main and obstructive right coronary artery disease), and 349 (10.9%) had evidence of prior revascularization.

Results were similar among those who underwent first coronary assessment concurrent with or after HF diagnosis (n=2,567): 933 (36.6%) had no CAD/negative stress, 707 (27.5%) had non-obstructive CAD, 867 (34.0%) had obstructive CAD or a positive stress test, and 60 (2.3%) were nondiagnostic or missing reports.

### Target Trial to Estimate Association with Composite Outcome & Mortality

Among 14,829 individuals included in the cohort, 5,972 met the inclusion criteria for the target trial of coronary assessment at the time of HF diagnosis (age<80, no prior coronary assessment, no urgent/emergent indication for coronary angiography, and no metastatic cancer or cirrhosis, and completed echocardiogram) including 627 who underwent testing and 5,345 who did not. Median follow up was 1725 days (IQR 617, 3206) among those who completed coronary assessment and 1317 days (IQR 390, 2805) among those who did not. Overall, 56% of the study population was diagnosed from 2002-2012 and 44% were diagnosed from 2013-2019, with a median follow-up time of 7.2 years in the earlier period and 2.1 years in the more recent period. At the end of follow-up, 201 (32.1%) who underwent early coronary assessment had died compared to 2,008 (37.6%) among those who did not (unadjusted p=0.007). For the primary composite outcome, 219 (34.9%) and 2,071 (38.8%), respectively, had died or underwent emergent coronary angiography (unadjusted p=0.06). Of eligible participants who did not undergo coronary assessment within 30 days, 639 (12%) crossed over and underwent coronary assessment at a median 380 days (IQR 116, 1090) after HF diagnosis.

Estimated probability of coronary assessment ranged from 3% in the lowest decile propensity score to 52% in the highest decile, so the propensity score successfully stratified probability of coronary assessment; the Hosmer-Lemshow test was consistent with adequate goodness of fit (p=0.70) and the area under the receiver operating curve was 0.75. All variables were well-balanced across propensity score quintiles.

Among eligible patients, elective coronary artery assessment at the time of HF diagnosis was associated with 16% lower risk of all-cause mortality over the entire study period (HR 0.84; 95% CI 0.72-0.98; p=0.025; Figure 3). Coronary artery assessment was associated with 14% lower risk for the composite outcome: HR 0.86 (95% CI 0.73-0.995; p=0.04). Estimates were unchanged in sensitivity analysis including race/ethnicity, unstable housing, medical history, and substance use for mortality (HR 0.85, 95% CI 0.73-0.99; p=0.04) and for the composite outcome (HR 0.87, 95% CI 0.75-1.02; p=0.08). Given possible violations of the assumption of proportional hazards (Supplemental Figure), we repeated the analyses truncating follow-up time at different intervals. Overall findings were robust to the censoring interval chosen (Supplemental Table 2).

**Figure 2.**
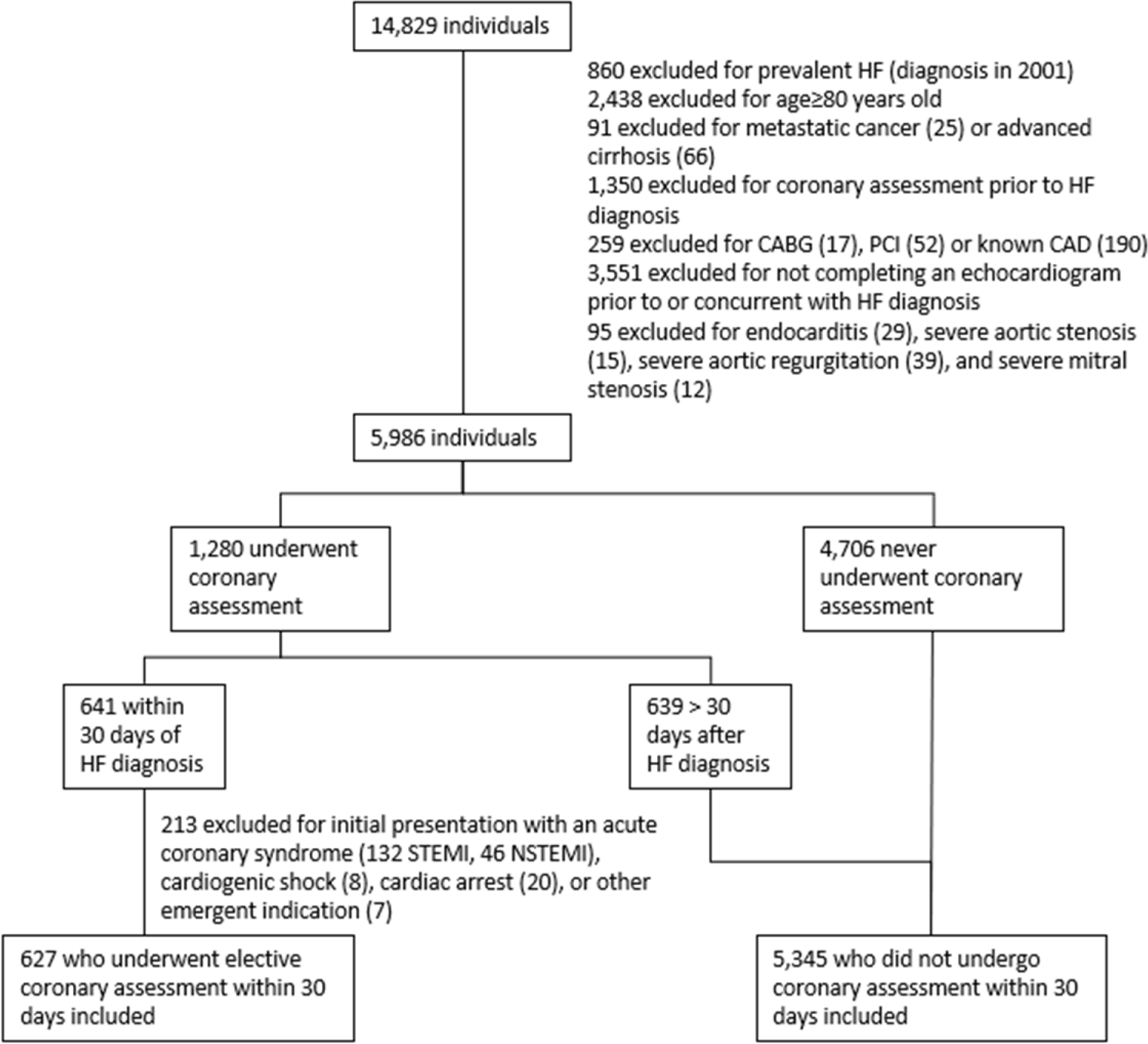
Consort Diagram for Target Trial of Elective Coronary Assessment at the Time of HF Diagnosis. To make good use of our observational data, we emulated a randomized controlled trial of elective coronary assessment at the time of heart failure diagnosis by creating a hypothetical trial of individuals “assigned” to early coronary assessment compared to those “assigned” to not undergo coronary assessment. This figure shows how we excluded individuals prevalent heart failure, with known CAD, competing diagnoses (cirrhosis/cancer), then by timing of coronary assessment, and finally excluding those whose initial presentation necessitated emergent coronary angiography (who would be more likely to benefit).

**Figure 3.**
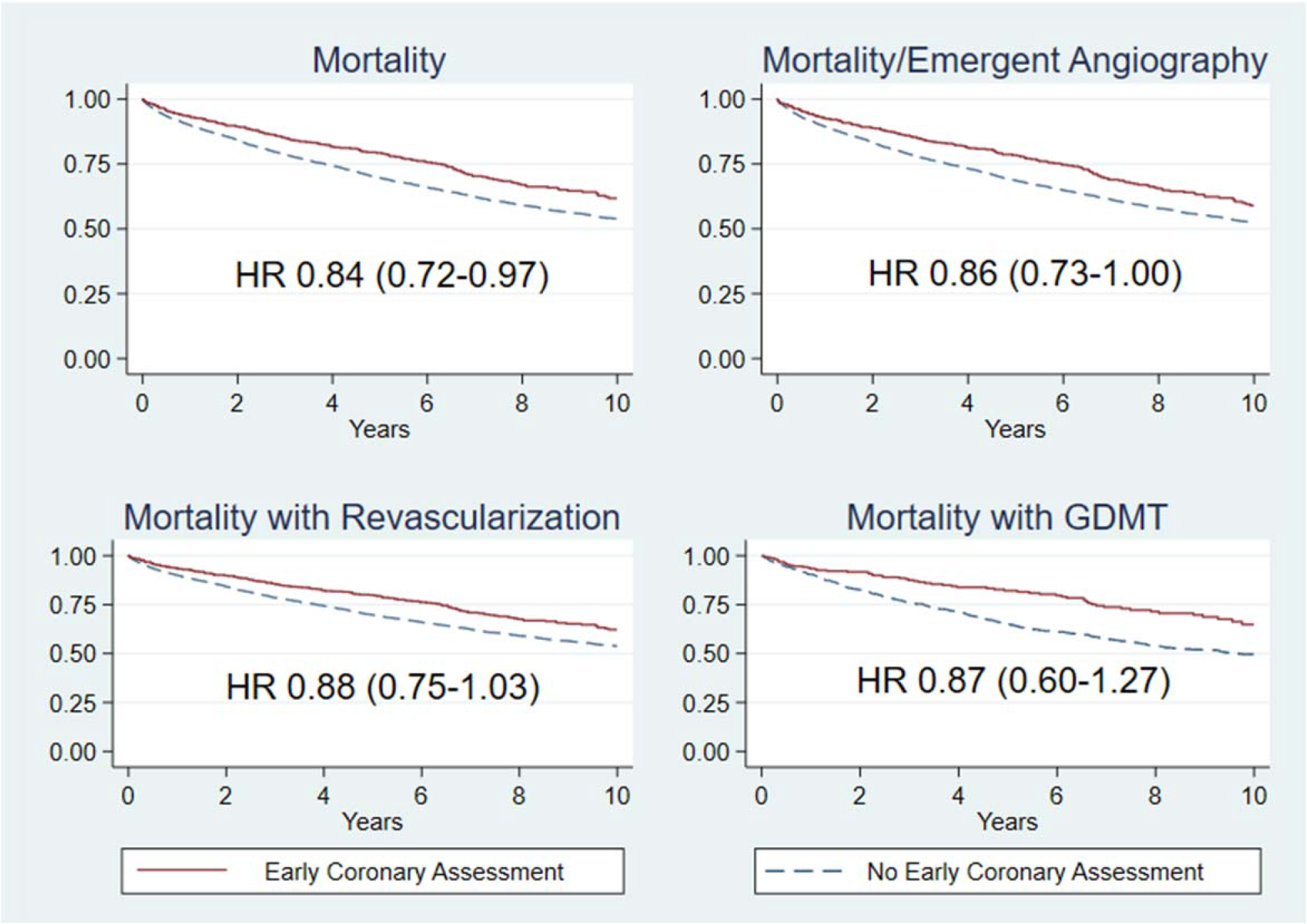
Association of Early Coronary Assessment in Heart Failure with Outcomes. After defining the “target trial” we adjusted for age, sex, propensity for coronary assessment as a restricted cubic spline, and HF hospitalization prior to testing and show the adjusted survival curves by concurrent coronary assessment. Hazard ratios are comparing those who completed early coronary assessment to those who did not, with the bottom two panels additionally adjusting for revascularization (bottom left) and use of GDMT among hospitalized patients with HFrEF (bottom right)

Accounting for age, sex, race, medical history, substance use, hospitalization at the time of diagnosis, HFrEF, and propensity for testing, early coronary assessment was associated with an absolute reduction in mortality of 4.4% (95%CI 0.6-8.3), or a number needed to test of 23 (95%CI 12-181) to save one life over a median follow-up of 3.8 years.

### Specific Subgroups of Interest: Female Patients, Hospitalized Patients, Those with HFrEF, Those Referred for “HF Workup,” and Those Found to have CAD

The effect estimates for coronary assessment on mortality did not vary by sex, with overall hazard ratios of 0.85 (95% CI 0.59-1.12) for women and 0.84 95% CI (0.71-1.00) for men (p_interaction_=0.86). Although not statistically significant, the effect estimates were stronger among those hospitalized at HF diagnosis (HR 0.72, 95% CI 0.55-0.93) compared to those not hospitalized (HR 0.92, 95% CI 0.76-1.11; p_interaction_=0.12). There was no difference by HFrEF (LVEF<40%) vs HFpEF: HR 0.84 in HFrEF (95% CI 0.69-1.01) vs HR 0.86 in HFpEF (0.66-1.11) (p_interaction_=0.89) or by regional wall motion abnormalities on the concurrent echocardiogram (p_interaction_=0.41). Among those who completed early coronary assessment, compared to no evidence of CAD or negative stress test, obstructive CAD was associated with higher risk (HR 1.30 95% CI 1.01-1.67). Among those who underwent coronary angiography, only multivessel CAD was associated with higher risk (HR 1.47, 95%CI 1.06-2.04).

### Role of Revascularization, GDMT & Outpatient Follow-up

We were able to ascertain revascularization records for 294/321 (92%) with obstructive CAD. Early coronary evaluation was associated with much higher likelihood of undergoing revascularization (11.2% vs 1.6%, p<0.001; adjusted OR 6.7; 95% CI 4.7-9.7). Among those revascularized, revascularization strategies were not significantly different between those who did or did not undergo early coronary assessment; 49% vs 40% received CABG (p=0.31) and 56 vs 62% received PCI (p=0.44), respectively. About half in each group with multivessel disease received revascularization (43% vs 53%; p=0.16) including 55% with left main disease and 50% with 3 or more obstructed vessels.

A much lower proportion in the early coronary assessment group were revascularized in the setting of an acute coronary syndrome (ACS) (11% vs 90%; p<0.001). The median time from HF diagnosis to revascularization was approximately 19 days compared to 1145 days (3.1 years) among those who did not undergo early coronary assessment (p<0.0001). Acknowledging confounding from referral bias and the benefit of revascularization in the setting of ACS, revascularization was associated with improved mortality in both groups (HR 0.58; 95%CI 0.39-0.87; p_interaction_=0.47). Accounting for revascularization did not change the overall effect estimate (HR 0.88; 95% CI 0.75-1.03). Among those who underwent early coronary assessment, compared to a reference of no CAD (all of whom did not undergo revascularization), having multivessel CAD without revascularization was associated with higher risk (HR 2.31 95%CI 1.42-3.76) whereas revascularized multivessel CAD was associated with lower risk (HR 0.47 95%CI 0.23-0.95) regardless of revascularization strategy with CABG or PCI. The extent to which the observed benefit is attributable to revascularization versus selection effects cannot be determined from our data, but the magnitude of the apparent benefit suggests confounding.

Among those who met the inclusion criteria for the target trial and were hospitalized, we considered whether ambulatory guideline directed medical therapy (GDMT) for HF and CAD could explain the apparent benefit of coronary assessment on mortality. To do this, we restricted our analysis to those hospitalized at the time of HF diagnosis for whom we had data on medical therapy and timing. For HF GDMT (beta blocker, ACE-I/ARB, and mineralocorticoid receptor antagonist) we only included those with HFrEF (n=791; 196 who underwent early coronary assessment), but for aspirin and statin we did not restrict by HF type (n=1104; 230 who underwent early coronary assessment).

A much higher proportion who underwent coronary assessment were prescribed each class of medical therapy prior to hospitalization compared to those who did not undergo coronary assessment (p<0.01 for each medication class, Table 4). Fewer than 10% of eligible individuals not already on therapy were initiated on each class of GDMT after hospitalization with no differences between groups. Prescriptions for aspirin, statin, ACE-I/ARB, beta-blocker, and mineralocorticoid receptor antagonist were each associated with lower hazard for mortality.

**Table 4.** Medical Therapy Among those Hospitalized for Heart Failure By Coronary Assessment.

|  | Timing | Coronary Assessment | No Coronary Assessment | Unadjusted p value | Adjusted HR for Mortality |
| --- | --- | --- | --- | --- | --- |
| ACE Inhibitor/<br>Angiotensin<br>Receptor Blocker | Pre-Hospitalization | 94 (48.0%) | 207 (34.8%) | <0.001 |  |
|  | Started | 13 (6.6%) | 27 (4.5%) |  |  |
|  | Ever | 107 (55.0%) | 234 (39.3%) | <0.001 | 0.40 (0.30, 0.53) |
| Beta Blocker | Pre-Hospitalization | 97 (49.5%) | 210 (35.3%) | 0.001 |  |
|  | Started | 12 (6.1%) | 32 (5.4%) |  |  |
|  | Ever | 109 (55.6%) | 242 (40.7%) | <0.001 | 0.39 (0.30, 0.52) |
| Mineralocorticoid<br>Receptor<br>Antagonist | Pre-Hospitalization | 46 (23.5%) | 91 (15.3%) | 0.009 |  |
|  | Started | 6 (3.1%) | 13 (2.2%) |  |  |
|  | Ever | 52 (26.5%) | 104 (17.5%) | 0.006 | 0.52 (0.35, 0.78) |
| Aspirin | Pre-Hospitalization | 78 (33.9%) | 197 (22.5%) | <0.001 |  |
|  | Started | 20 (8.7%) | 63 (7.2%) |  |  |
|  | Ever | 98 (42.6%) | 260 (29.7%) | <0.001 | 0.48 (0.38, 0.61) |
| Statin | Pre-Hospitalization | 87 (37.8%) | 194 (22.2%) | <0.001 |  |
|  | Started | 16 (7.0%) | 42 (4.8%) |  |  |
|  | Ever | 103 (44.8%) | 236 (27.0%) | <0.001 | 0.45 (0.35, 0.58) |

Table Legend: Individuals with HFrEF hospitalized prior to coronary assessment were included for the beta-blocker, ACE/ARB, and MRA analysis (n=791; 196 who underwent coronary assessment at time of HF diagnosis and 595 who did not). Individuals hospitalized at the time of diagnosis prior to coronary assessment regardless of HFrEF were included for the aspirin/statin analyses (n=1104; 230 who underwent coronary assessment and 874 who did not). Adjusted Cox Proportional Hazards models include age, sex, hypertension, diabetes, chronic kidney disease, coronary assessment, cubic spline for propensity for testing, and each individual class of medical therapy.

Among those with HFrEF hospitalized for HF, undergoing early coronary assessment was associated with higher odds of ever being prescribed HF GDMT (59% vs 43%; adjusted OR 2.5; 95%CI 1.7-3.6), but not with greater initiation of HF GDMT (11.1% vs 7.9%; adjusted OR 1.4; 95%CI 0.8-2.5). Being on at least one class of GDMT was associated with a 57% lower hazard for mortality (HR 0.43; 95% CI 0.33-0.57). Accounting for the number of GDMT classes prescribed, the effect of coronary assessment was similar (HR 0.87; 95% CI 0.60-1.27; p_interaction_=0.88). Because GDMT was prescribed at much higher rates from 2013 onward (74% vs 26%, p<0.001), we subsequently restricted the analysis to only those diagnosed in 2013 and later. Among those diagnosed in 2013 and later, there was no benefit of elective coronary assessment (HR 1.02 95% CI 0.75-1.39), but the median follow-up time was only 2 years for this subset as compared to 7 years among those diagnosed 2002-2012, and the interaction term was not statistically significant (p_interaction_=0.15). Accounting for the annual proportion with hospitalized HFrEF receiving outpatient GDMT did not attenuate the overall benefit (HR 0.84; 95% CI 0.72-0.98).

Among those admitted with HF at the time of diagnosis, completing coronary assessment was associated with higher odds of attending outpatient follow-up within 30 days (83% vs 63%, p<0.001) which was in turn associated with lower mortality (HR 0.81 95% CI 0.68-0.95), but there was no effect modification of outpatient follow-up status on the benefit of completing coronary assessment (p_interaction_=0.91).

### Sensitivity Analyses to Check Robustness of Findings Including Use of Inverse Probability of Treatment Weighting and Nearest-Neighbor Matching as Alternative Approaches

To check the robustness of our criteria for excluding those with acute myocardial infarction and other indications for coronary assessment, when we limited the coronary assessment group to the 303 whose documented indication for coronary assessment was “cardiomyopathy workup” or equivalent, results were similar: HR 0.82 (95% CI 0.66-1.01).

To check if our results were sensitive to our a priori analytic choice of using the restricted cubic spline of the propensity score for coronary assessment rather than inverse probability of treatment weighting (IPTW), we repeated our analysis using IPTW focusing on 4-year mortality and overall mortality. With this approach, coronary assessment was associated with reduced mortality at 4 years (OR 0.54; 95% CI 0.40-0.74) and overall (OR 0.64; 95% CI 0.50-0.83), similar to the results from the propensity adjusted analysis (OR 0.66 at 4 years and 0.81 overall).

Using IPTW, the average treatment effect over the whole study was a 9.6% absolute reduction in mortality (95% CI 4.5-14.7) or 6.6 at 4 years (95%CI 3.2-10.0). We replicated these findings using a nearest-neighbor matching approach with an average treatment effect of 6.8% absolute reduction in mortality (3.1-10.6) over 4 years and 3.3% (−1.1 to 7.9%) overall.

## Discussion

In this study of nearly 15,000 individuals with HF from 2001-2019 who received care in the municipal safety-net system in San Francisco, we found significant differences in who receives coronary artery assessment that did not align with risk of CAD or risks of coronary assessment. Our second question was whether this matters for clinical outcomes including mortality and a composite of mortality and emergent coronary angiography. Our “target trial” emulating a randomized trial of elective coronary assessment after HF diagnosis suggested a statistically and clinically significant improvement in mortality and in the composite outcome with elective coronary angiography. Individuals who received early coronary assessment were much more likely to be revascularized, but also had higher rates of GDMT prescription prior to testing. Although there is a risk of residual confounding from our use of observational, EHR-collected data, our findings suggest that that early coronary assessment may be beneficial.

### Patterns in Coronary Artery Assessment

Within a safety-net setting, fewer than one in five had coronary assessment concurrent with or after heart failure diagnosis. We found that women, older individuals, and those with documented unstable housing were less likely to complete coronary assessment, as well as differences by race and ethnicity, past medical history, and substance use. These patterns were not explained by coronary risk: for example, diabetes and HIV were associated with lower odds of testing even accounting for chronic kidney disease. Our findings are similar to several recent studies that have found a low proportion of individuals who underwent coronary assessment among individuals with incident heart failure without known CAD ranging from 20% in Denmark,^13^ 17.5% at index hospitalization and 27.4% within 90 days among commercially insured and Medicare patients in the United States,^14^ and 40% among those hospitalized with HFrEF who survived 90 days without rehospitalization in the Veteran’s Affairs system in the United States.^15^ Similarly, among those with incident heart failure in the United States, 35% underwent coronary assessment within 90 days of heart failure diagnosis, with similar patterns as found in our study, with younger, male, hospitalized patients, with a lower ejection fraction more likely to have coronary assessment.^10^ These patterns, particularly lower testing among women even accounting for differences in risk by age, by race and ethnicity, and ejection fraction may be attributable to biases in referral (provider-level sexism and racism), differences in resources available to complete testing (structural sexism and racism), or differences in acceptability of testing (patient-physician trust and patient preferences). Not surprisingly, those referred for early coronary assessment were much more likely to ever undergo revascularization and went revascularization sooner after HF diagnosis, with similar rates of CABG and PCI.

### Coronary Assessment and Outcomes

There are no randomized controlled studies of coronary assessment in heart failure, and we reproduced the findings from earlier observational studies that suggested coronary assessment may be associated with improved outcomes. An analysis from the OPTIMIZE-HF study, which found that coronary assessment was associated with greater use of GDMT and improved outcomes among those found to have significant CAD, did not account for propensity for referral for coronary assessment and stratified based on nonischemic and ischemic cardiomyopathy which is unknown without coronary assessment.^8^ Similarly, an observational study within the Veterans Affairs system found that ischemic evaluation was associated with reduced mortality and higher use of GDMT, but that study excluded those who did not survive more than 90 days, creating immortal time bias and excluding immediate harms from invasive coronary assessment.^15^ Observational data from Ontario suggest that early invasive coronary angiography within 2 weeks of HF diagnosis is associated with four times higher rates of revascularization within 90 days, 26% lower morality and 16% lower heart failure readmissions at two years.^9^ However, that study included those with known CAD and those presenting with acute coronary syndromes who are much more likely to benefit from immediate invasive angiography. Not surprisingly, we also found that early coronary assessment was associated with higher GDMT prescriptions (even prior to coronary assessment) and higher likelihood of revascularization.

We used a rigorous approach to use observational data to emulate a randomized controlled trial, including creating robust inclusion and exclusion criteria to restrict inclusion to those with equipoise regarding coronary assessment, starting follow up time for all individuals at the time of incident HF diagnosis, limiting coronary assessment to a 30-day grace period to minimize immortal time bias, using propensity scores to adjust for confounders measured at the time of study eligibility, and conducting sensitivity analyses. Our results were robust to our analytic assumptions using alternative approaches. Including individuals who eventually underwent coronary assessment after 30 days in the “no coronary assessment group” is analogous to cross-over in a randomized trial; crossover would tend to bias our results toward the null, but this approach is the best approximation to the intention to treat approach. None of the subgroups of interest met our specified criteria for statistical significance but there were nonsignificant trends toward greater benefit among those hospitalized at the time of their heart failure diagnosis, those with HFrEF, and those with regional wall motion abnormalities on echocardiogram.

### Future Directions

Our study leaves several important questions unanswered. First, does the benefit we found persist in an era of widespread use of contemporary GDMT for HF with angiotensin receptor blockers/neprilysin inhibitors (ARNI), beta-blockers, mineralocorticoid receptor antagonists, and sodium glucose cotransporter-2 inhibitors (SGLT2i)? Our study suggests that if there is a benefit, participants will need to be followed for longer than 2 years to have any potential benefit outweigh the upfront risks from coronary assessment and subsequent revascularization among those found to have multivessel CAD.

Secondly, the role of coronary assessment in HF is most often linked to identifying patients who may benefit from revascularization, and we found that early coronary assessment was associated with much higher likelihood of ever undergoing revascularization and earlier revascularization. The STICHES trial demonstrated that surgical revascularization for ischemic cardiomyopathy is associated with improvements in long term outcomes including mortality and rehospitalization at 10 years,^16^ but did not demonstrate a statistically significant result at 5 years in an earlier era of medical therapy. Results from two large randomized controlled trials, ISCHEMIA^6^ (which excluded patients with LVEF<40%) and REVIVED-BCIS2 (which only included patients with LVEF<35%),^7^ have called into question the role of revascularization in chronic stable angina and ischemic cardiomyopathy, respectively. Even with these studies (with only medium-term results reported so far), there are unanswered questions including whether percutaneous coronary intervention and CABG should be considered equivalent in this setting, the role for viability or functional testing, and most importantly whether revascularization improves HF symptoms, quality of life, and long-term mortality.

To our knowledge, there are no randomized trials of the role of coronary assessment in HF and there is limited evidence to guide whom should have their coronaries assessed after diagnosis with HF, best strategies for initial test selection, and ultimately whether early elective coronary assessment among patients with new HF prospectively improves patient-centered outcomes. To definitively answer these questions may require a pragmatic randomized clinical trial embedded in routine clinical care, especially given the consistently low rates of referral for coronary assessment across the published studies.

### Limitations

The first limitation is that this is an observational study based primarily on use of electronic health records. Use of ICD codes to ascertain propensity for coronary assessment results in a meaningfully high risk of residual confounding, as many clinical and socioeconomic factors are not well-captured in the electronic records. Secondly, we were unable to use an intention-to-treat approach as we were not able to ascertain those referred for testing who did not complete it. Those who complete coronary artery testing are more likely to attend outpatient follow up, take prescribed medications, and undergo revascularization; limiting our study population to those who had completed an echocardiogram only partially accounts for this selection bias. A third limitation is that we only included coronary assessment performed within SFHN or ordered by SFHN and performed at UCSF Health (nuclear stress), which would tend to bias the results toward the null, although we were able to ascertain revascularization outcomes across the major regional health systems due to EHR connectivity. Although we planned to estimate atherosclerotic cardiovascular risk using the pooled cohort equations as a proxy for who should be referred for coronary assessment, ultimately, we did not do this as hospital blood pressures may reflect acute illness, medication use, lipid panels were missing for many individuals, and we could not verify current vs past smoking. We also did not have access to time-varying covariates except outpatient prescription data which was only available for those who were hospitalized before and after hospitalization, limiting our ability to adjust for the time-varying nature of GDMT in the whole study population. Finally, this study was conducted prior to the use of SGLT2i and widespread ARNI use. Despite our best efforts to emulate a target trial, these issues make the interpretation of our findings less conclusive despite the robustness to our assumptions.

## Conclusions

Among individuals with HF in a safety net setting, we found significant differences in who completed coronary assessment that are not explained by coronary risk factors. Our target trial emulation suggests early elective coronary assessment among patients with HF without another indication for urgent coronary assessment is associated with improved mortality, more revascularization, and higher use of HF GDMT in a safety net population. The extent to which our findings reflect a true benefit from early coronary assessment rather than unmeasured confounding from selection effects remains uncertain, suggesting that this clinical question requires a randomized clinical trial to answer with confidence.

## Data Availability

The deidentified dataset will be made available upon reasonable request to the corresponding author.

